# Robust use of phenotypic heterogeneity at drug target genes for mechanistic insights: application of *cis*-multivariable Mendelian randomization to *GLP1R* gene region

**DOI:** 10.1101/2023.07.20.23292958

**Authors:** Ashish Patel, Dipender Gill, Dmitry Shungin, Christos S. Mantzoros, Lotte Bjerre Knudsen, Jack Bowden, Stephen Burgess

**Affiliations:** MRC Biostatistics Unit, University of Cambridge, UK; Department of Epidemiology and Biostatistics, School of Public Health, Imperial College London, UK; Chief Scientific Advisor Office, Research and Early Development, Novo Nordisk, Denmark; Human Genetics Centre of Excellence, AI and Digital Research, Novo Nordisk, Denmark; Department of Internal Medicine, Beth Israel Deaconess Medical Center, Harvard Medical School, USA; Department of Internal Medicine, Boston VA Healthcare System, Harvard Medical School, USA; Department of Clinical and Biomedical Sciences, University of Exeter, UK; Department of Genetics, Novo Nordisk Research Centre Oxford, U.K; Cardiovascular Epidemiology Unit, University of Cambridge, UK

**Author notes:** **Conflict of Interest Disclosure:** D.G. and J.B. are part time employees of Novo Nordisk. D.S. and L.B.K. are full time employees of Novo Nordisk.

**Keywords:** drug target Mendelian randomization, phenotypic heterogeneity, overdispersion heterogeneity, GLP1R agonists

## Abstract

Phenotypic heterogeneity at genomic loci encoding drug targets can be exploited by multivariable Mendelian randomization to provide insight on the pathways by which pharmacological interventions may affect disease risk. However, statistical inference in such investigations may be poor if overdispersion heterogeneity in measured genetic associations is unaccounted for. In this work, we first develop conditional F-statistics for dimension-reduced genetic associations that enable more accurate measurement of phenotypic heterogeneity. We then develop a novel extension for two-sample multivariable Mendelian randomization that accounts for overdispersion heterogeneity in dimension-reduced genetic associations. Our empirical focus is to use genetic variants in the *GLP1R* gene region to understand the mechanism by which GLP1R agonism affects coronary artery disease (CAD) risk. Colocalization analyses indicate that distinct variants in the *GLP1R* gene region are associated with body mass index and type 2 diabetes. Multivariable Mendelian randomization analyses that were corrected for overdispersion heterogeneity suggest that bodyweight lowering rather than type 2 diabetes liability lowering effects of GLP1R agonism are more likely contributing to reduced CAD risk. Tissue-specific analyses prioritised brain tissue as the most likely to be relevant for CAD risk, of the tissues considered. We hope the multivariable Mendelian randomization approach illustrated here is widely applicable to better understand mechanisms linking drug targets to diseases outcomes, and hence to guide drug development efforts.

## Introduction

By *phenotypic heterogeneity*, we refer to differences in the effects of distinct genetic variants in a single gene region on observable traits (Nussbaum et al., 2007). The increasing size and scope of genome-wide association studies has resulted in the discovery of multiple causal variants for many gene regions (Visscher et al., 2017; Pasaniuc and Price, 2017; Yang et al., 2012). In some cases, different causal variants in the same gene have distinct patterns of association with gene expression, molecular, and phenotypic traits. The principle of Mendelian randomization is that genetic variants can be used as unconfounded proxies to understand the consequences of intervening on the pathway or trait affected by the variants (Davey Smith and Hemani, 2014). The use of genetic associations of variants in a genetic region relating to a molecular target to make causal inferences is known as *cis*-Mendelian randomization (Schmidt et al., 2020).

Genes code for proteins, which make up the majority of drug targets, particularly for small molecule and biologic compounds. Previous work has demonstrated that *cis*-Mendelian randomization focused on genes coding for drug target proteins can be used to anticipate the effects of their pharmacological perturbation (Gill et al., 2021; Holmes et al., 2021; Daghlas and Gill, 2023). The human relevance of such insights offers considerable advantages over animal models, which can be difficult to translate to patient populations (Hingorani et al., 2019). Further, the random allocation of genetic variants at conception means that their associations with clinical traits are less vulnerable to the confounding from environmental factors and reverse causation bias that can hinder causal inference in traditional epidemiological study designs (Burgess et al., 2023). However, it is also important to appreciate that genetic variants mimicking drug class effects typically inform on small, lifelong perturbations of the corresponding target, which contrasts with the effects of discrete interventions of larger magnitude that are most commonly encountered in clinical practice (Gill et al., 2021). As such, it is not advisable to directly translate effect estimates from genetic analyses to those that might be expected in clinical practice.

Multivariable Mendelian randomization uses shared genetic predictors of related traits to distinguish between the causal effects of the traits (Sanderson et al., 2019). Such analyses have the potential to provide important mechanistic insights to identify causal risk factors in a way that univariable Mendelian randomization analyses do not: univariable analyses may falsely conclude that a non-causal trait is causal because it is genetically correlated with a causal trait. Multivariable *cis*-Mendelian randomization studies can therefore provide evidence on the causal mechanism linking the gene to the outcome that could guide the design of a pharmacological intervention trial.

There are, however, several methodological difficulties of multivariable *cis*-Mendelian ran-domization. First, in order to identify multivariable trait effects, we require genetic predictors of each trait that are not collinear. This is potentially problematic because it is often not possible to conduct such analyses using a set of variants pruned to minimal correlation since such variants are likely to be few in number and unlikely to provide enough information to reliably estimate several trait effects (Batool et al., 2022). Conversely, the challenge with using a larger number of correlated variants in a single gene region is that they are typically highly correlated. Hence, even if different causal variants have different mechanistic consequences, disentangling the effects of these mechanisms on a disease outcome is tricky.

Second, Mendelian randomization analyses must typically rely on summarised data, representing genetic associations (beta-coefficients and standard errors) from regression on the trait of each genetic variant in turn (Burgess et al., 2013). Modelling approaches using marginal genetic association estimates can suffer from numerical instability, as the matrix of correlations between genetic variants can be near singular (ill-conditioned) even if pairwise correlations between the genetic variants are pruned at a threshold level (Zou et al., 2022).

Previous investigations have tackled these two challenges by using dimension reduction techniques to improve the stability of summary data analyses with a large number of correlated variants, including principal component analysis (Burgess et al., 2017) and factor analysis (Patel et al., 2023). However, these methods may be vulnerable to an important practical problem of *overdispersion heterogeneity* in variant–outcome associations, whereby genetic variants have direct effects on the outcome, not via their effects on included traits, which manifests as heterogeneity in a random-effects model. The failure to account for overdispersion heterogeneity may lead to overly precise confidence intervals for estimated trait effects and thus misleading analyses, as previously shown for univariable Mendelian randomization (Zhao et al., 2020).

We emphasise that the two types of heterogeneity discussed in this paper have different consequences: phenotypic heterogeneity enables us to reliably estimate multivariable trait effects based on a single gene region, whereas overdispersion heterogeneity presents a challenge for inference using these estimated trait effects. Therefore, following previous terminology (Sanderson et al., 2019), we can think of phenotypic heterogeneity as a ‘good’ type of heterogeneity, and overdispersion heterogeneity as a ‘bad’ type for multivariable Mendelian randomization estimation.

In this work, we bridge this methodological gap by providing a way forward for summary data multivariable *cis*-Mendelian randomization analysis that is robust to overdispersion heterogeneity. Our empirical focus concerns GLP1R agonists, which have proven effective for the treatment of type 2 diabetes and obesity (Aroda et al., 2019; Wilding et al., 2021), and for preventing cardiovascular events in people with type 2 diabetes (Marso et al., 2016; Gerstein et al., 2019; Hernandez et al., 2018), with a cardiovascular outcome clinical trial underway in people with obesity (Ryan et al., 2020). We perform multivariable *cis*-Mendelian randomization for variants in the *GLP1R* gene region: first, considering body mass index (BMI) and type 2 diabetes (T2D) as distinct risk factors, representing two outcomes affected by GLP1R agonism; and second, considering *GLP1R* gene expression in different tissues as risk factors. In each case, the outcome is coronary artery disease (CAD) risk. The aim of these analyses is to better understand the biological pathway by which GLP1R agonism affects CAD risk. These risk factors represent pathways by which the effect of GLP1R agonism may affect CAD risk, rather than the totality of the effect of the risk factor itself. Software code to implement the methods introduced here are available in the MendelianRandomization R package (Yavorska et al., 2020).

## Methods

We perform a simulation study and an empirical investigation. The empirical investigation has three connected elements. First, we assess whether genetic associations with BMI and T2D at the *GLP1R* gene region colocalize, and we compute conditional F-test statistics for dimension-reduced genetic associations. Second, we perform multivariable *cis*-Mendelian randomization analyses considering the effects of BMI and T2D liability on CAD risk based on variants in the *GLP1R* gene region. Third, we perform multivariable *cis*-Mendelian randomization analyses considering the effects of *GLP1R* gene expression on CAD risk based on variants in the *GLP1R* gene region.

The first analysis explores whether there is phenotypic heterogeneity for BMI and T2D at the *GLP1R* locus, to verify that their genetically-predicted effects on CAD risk can be reliably estimated. The second analysis explores whether any effect of perturbing *GLP1R* pathways on CAD risk is due to the effect of BMI or T2D liability. The third analysis explores the likely tissue at which the effect of *GLP1R* perturbation on CAD risk occurs. A schematic diagram illustrating these elements is presented as Figure 1.

**Figure 1.**
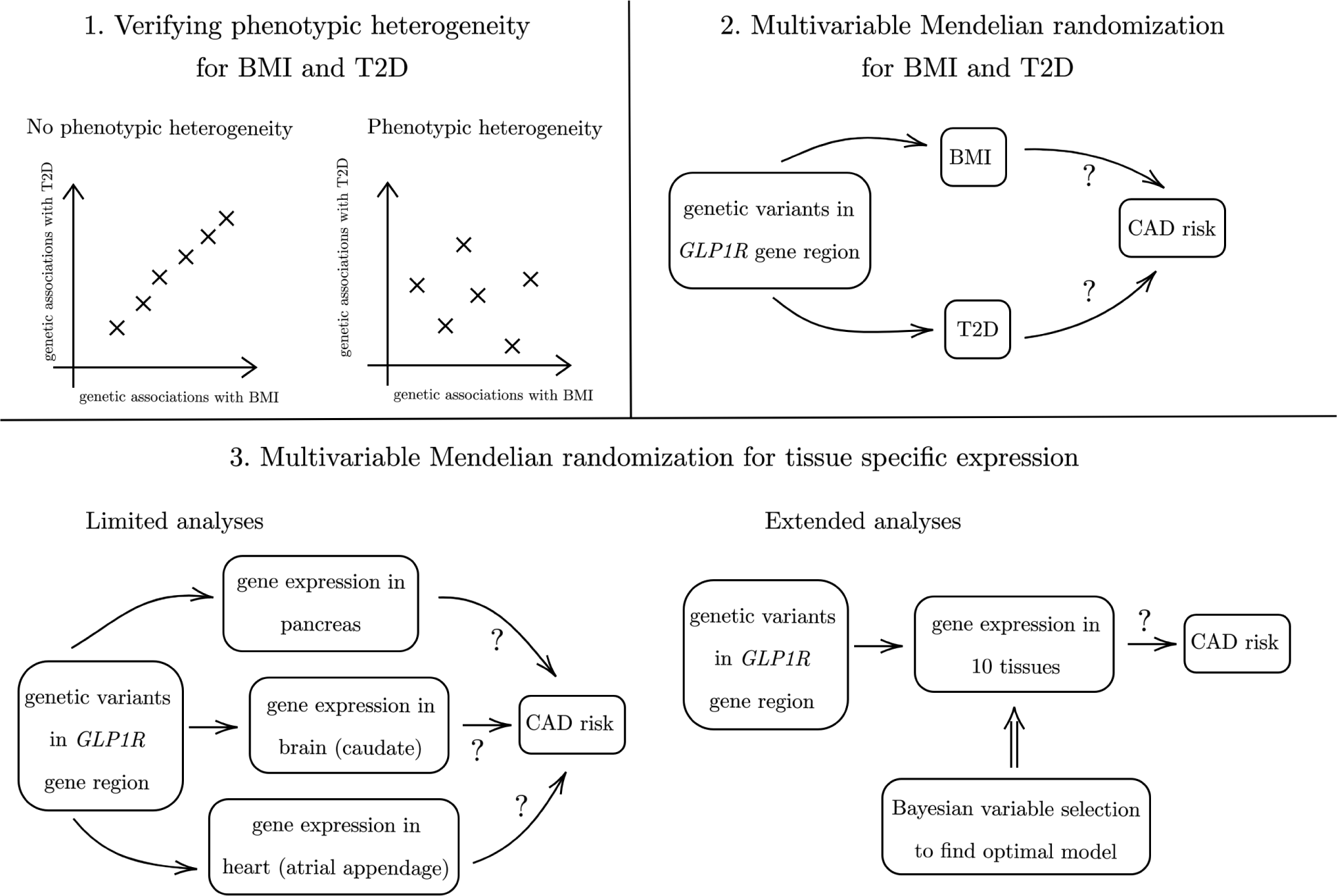
Schematic diagram of our three connected analyses.

### Data sources

Genetic associations with BMI were obtained from a meta-analysis of 694,649 individuals of European ancestries from UK Biobank and the Genetic Investigation of ANthropometric Traits (GIANT) consortium (Pulit et al., 2019). Genetic associations with T2D risk were obtained from a meta-analysis of 80,154 cases and 853,816 controls of European ancestries from the DIAbetes Meta-ANalysis of Trans-Ethnic association studies (DIAMANTE) (Mahajan et al., 2022) consortium. Genetic associations with gene expression were estimated in 838 participants from the Genotype-Tissue Expression (GTEx) project version 8 (GTEx Consortium, 2020). Genetic associations with CAD were obtained from a GWAS of 1,165,690 individuals (181,522 cases) mostly of European ancestries from the Coronary ARtery DIsease Genome wide Replication and Meta-analysis plus The Coronary Artery Disease Genetics (CARDIoGRAMplusC4D) consortium (Aragam et al., 2022). Correlation matrices for variants were estimated using 367,703 unrelated UK Biobank participants of European ancestries (Astle et al., 2016). We considered associations of genetic variants in a region ± 100kbp of the *GLP1R* gene (chr6:39,016,557-39,059,079 in GRCh37/hg19).

We perform two tissue-specific analyses: a limited analysis, comparing gene expression in three tissues; and an extended analysis, comparing gene expression in ten tissues. The three tissues in the limited analysis were chosen as those with high levels of *GLP1R* expression and in biologically plausible tissues: brain, pancreas, and heart. The tissue subtype that had greatest *GLP1R* expression: brain-caudate, pancreas, and heart-atrial appendage were used in the analysis. The ten tissues in the extended analysis were chosen based on *GLP1R* expression levels alone, and additionally include thyroid, testis, stomach, nerve, lung, heart-left ventricle, and brain-hypothalamus.

### Statistical methods

Colocalization analyses may be used to assess whether two traits have distinct causal variants in a single genetic region, and thus can provide evidence for phenotypic heterogeneity. Colocalization was performed by the coloc method using the default prior settings (Giambartolomei et al., 2014; *p*_1_ = *p*_2_ = 10*^−^*^4^, *p*_12_ = 10*^−^*^5^), where *p_k_* is the prior probability that a genetic variant is associated with trait *k*, (*k* = 1, 2). We also performed a sensitivity analysis varying the value of *p*_12_, which represents the prior probability of a variant being causal for both traits.

Conditional F-statistics are an alternative way to assess phenotypic heterogeneity, which is *required* to reliably estimate risk factor effects on the outcome using multivariable Mendelian randomization. More formally, conditional F-statistics (Sanderson and Windmeijer, 2016; Sanderson et al., 2021) can be used to test for evidence against a rank reduction of one for any given risk factor, where the genetically-predicted effects of the risk factor can be expressed as a linear combination of other genetically-predicted risk factor effects (top-left panel of Figure 1). We computed conditional F-statistics for BMI and T2D; a higher value of the conditional F-statistic suggests greater evidence of phenotypic heterogeneity for that risk factor.

Multivariable *cis*-Mendelian randomization analyses were performed using the Principal Component analysis-based Generalised Method of Moments (PC-GMM) method. This method extends the previously published multivariable inverse-variance weighted principal component analysis method (Batool et al., 2022) in two directions: firstly, it uses the continuously updating generalised method of methods (GMM) method (Hansen et al., 1996), which is known to be less sensitive to weak instruments than other instrumental variable methods (Antoine and Renault, 2009; Chao and Swanson, 2005); and secondly, it can allow for heterogeneity in the model by incorporating an overdispersion parameter.

The intuition behind the approach is that the additional uncertainty due to overdispersion heterogeneity affects the variance of estimates rather than the bias. Hence, consistent estimation is possible without accounting for possible overdispersion, which allows us to propose an overdispersion parameter that can be used to correct standard errors. We refer to the method with an overdispersion parameter as “robust PC-GMM”.

For the unrobust version of the PC-GMM method that assumes there is no overdispersion heterogeneity, an overidentification test (OID test; Hansen, 1982) (henceforth, heterogeneity test) can be used to assess heterogeneity that is unexplained by the model; a rejection of this test indicates model misspecification, which is a sign of possible pleiotropy. Further technical details of conditional F-statistics and the robust PC-GMM method are found in Supplementary Material.

Comparison of models for the extended analysis was performed using the Mendelian Randomization Bayesian Model Averaging (MR-BMA) method (Zuber et al., 2020). Fitting a model with all ten tissues as risk factors can result in imprecise estimates that are difficult to interpret due to multicollinearity. Instead, the MR-BMA method fits models with each risk factor in turn, all pairs of risk factors, all triples of risk factors, and so on. Each model, representing a particular combination of risk factors, receives a posterior model probability based on its goodness-of-fit; the model that best explains the genetic associations with the outcome will receive the greatest posterior probability. Additionally, each risk factor is assigned a marginal inclusion probability, calculated as the sum of the posterior model probabilities for all models including that risk factor. The method is implemented using stochastic search, with a prior probability of *p* = 0.1 for each tissue, representing a prior expectation that one tissue is the true causal tissue.

The PC-GMM method requires an input of a risk factor correlation matrix. For our analysis of tissue-specific effects, only the trait correlations between the two brain tissues and between the two heart tissues were set at 0.7. Otherwise, the correlation entries were set equal to 0. Sensitivity to these values was investigated. Unless otherwise stated, all Mendelian randomization estimates are expressed as log odds ratios per 1 standard deviation increase in genetically-predicted levels of the risk factor.

### Simulation study

Our simulation study comprises two parts. The first part investigates how the bias of Mendelian randomization estimates varies with conditional F-statistics, a measure of phenotypic heterogeneity *ξ*. The second part investigates the ability of the robust PC-GMM method to provide reliable inferences under overdispersion heterogeneity *κ*^2^. An illustration of phenotypic and overdispersion heterogeneity in our simulation design is given in Figure 2.

**Figure 2.**
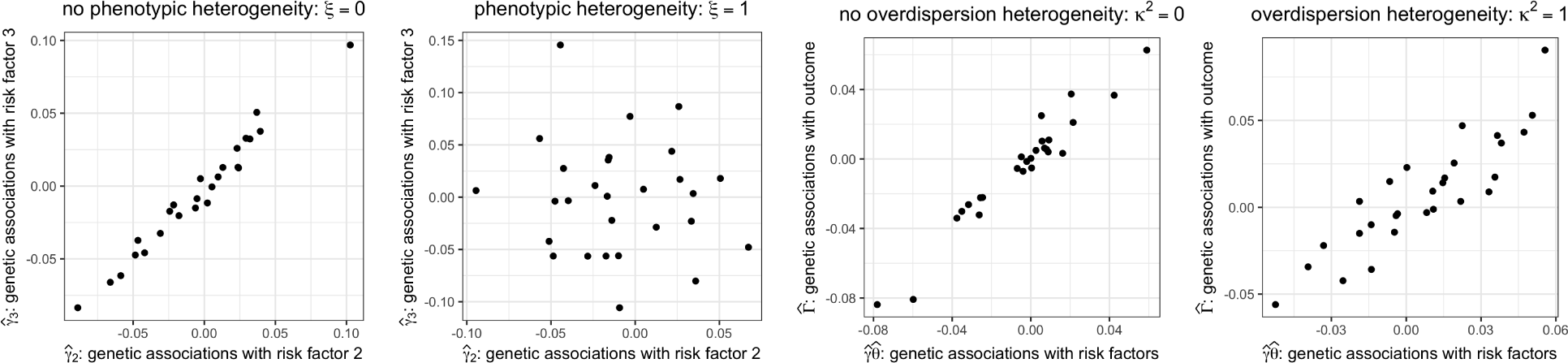
Simulated dimension-reduced genetic associations varying with phenotypic and overdispersion heterogeneity.

We generated two-sample summary data from a linear instrumental variable model with 3 risk factors and 200 instruments. Out of the 200 instruments, 15 instruments that were mutually weakly correlated (*R*^2^ *≤* 0.2) had non-zero fixed effects on at least one risk factor. The remaining 185 instruments had no effect on any risk factor. The instruments, *Z* = (*Z*_1_*,…, Z*_200_)*^t^*, were normally distributed with mean zero and were mutually correlated according to a measured genetic variant correlation matrix of the *GLP1R* gene region based on individuals of European ancestries (Supplementary Figure S1). The three risk factors were generated as 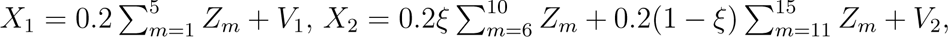, and 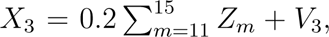, where the errors (*V*_1_*, V*_2_*, V*_3_) were normally distributed such that *var*(*V*_1_) = *var*(*V*_2_) = *var*(*V*_3_) = 1, *cor*(*V*_1_*, V*_2_) = *cor*(*V*_1_*, V*_3_) = *cor*(*V*_2_*, V*_3_) = 0.3. Hence, the parameter *ξ ∈* [0, 1] is a measure of phenotypic heterogeneity, with *ξ* = 0 implying no phenotypic heterogeneity for risk factors 2 and 3, and *ξ* = 1 implying phenotypic heterogeneity for all risk factors.

The sample sizes *n_X_* and *n_Y_* used to compute two-sample summary data were varied from 500 to 20, 000. Knowledge of the true instrument correlation matrix and a sample correlation matrix of risk factors was assumed. The impact of mis-specifying risk factor correlations is discussed in Supplementary Material. The number of principal components was chosen to explain 99.99% of variation of a sample weighted instrument correlation matrix. Each of the 200 instruments had a direct effect on the outcome which was normally distributed with mean zero and variance proportional to an overdispersion parameter *κ*^2^ ranging between 0 and 1. The outcome was generated as *Y* = *θ*_1_*X*_1_ + *θ*_2_*X*_2_ + *θ*_3_*X*_3_ + *Z*’α + *U*, where *U* is a mean zero error term with variance 1, and is correlated with the errors in the exposure model such that *cor*(*V_k_, U*) = 0.2 for *k* = 1, 2, 3. The random effect *α* was normally distributed with mean zero and variance equal to 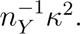. The causal effect of the 3 risk factors on the outcome was set equal to *θ* = (*θ*_1_*, θ*_2_*, θ*_3_)*^t^* = (*−*1*/*3, 0, 1*/*3)*^t^*. Therefore, for risk factors 1 and 3 we tested the ability of the robust PC-GMM method to successfully reject the null hypothesis of no causal effect, whereas for risk factor 2 we tested the ability of our method to control the type I error rate.

## Results

### Simulation results

The results of robust PC-GMM estimation and inference for the case of no overdispersion heterogeneity (*κ*^2^ = 0) and varying phenotypic heterogeneity (0 *≤ ξ ≤* 1) are displayed in Figure 3. The first row of Figure 3 shows that for large enough sample sizes the type I error rates of conditional F-tests were controlled at the nominal 5% level when *ξ* = 0, and the tests were able to detect phenotypic heterogeneity when *ξ /*= 0. The second and third rows of Figure 3 highlight that the estimates 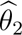 and 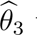 were heavily biased when the conditional F-statistics *F*_2_*_|−_*_2_ and *F*_3_*_|−_*_3_ were low; this estimation bias disappears when *ξ* is further away from 0 and when sample sizes are large (and hence the conditional F-statistics are larger). The final row of Figure 3 shows that the type I error rate for the test *H*_0_: *θ*_2_ = 0 was not controlled at 5% unless there was sufficient phenotypic heterogeneity and a large enough sample size. Similarly, the power to detect the non-zero effect *θ*_3_ = 1/3 was higher for larger values of the conditional F-statistic, since the estimate *θ*_3_was otherwise biased toward *θ*_2_ = 0.

**Figure 3.**
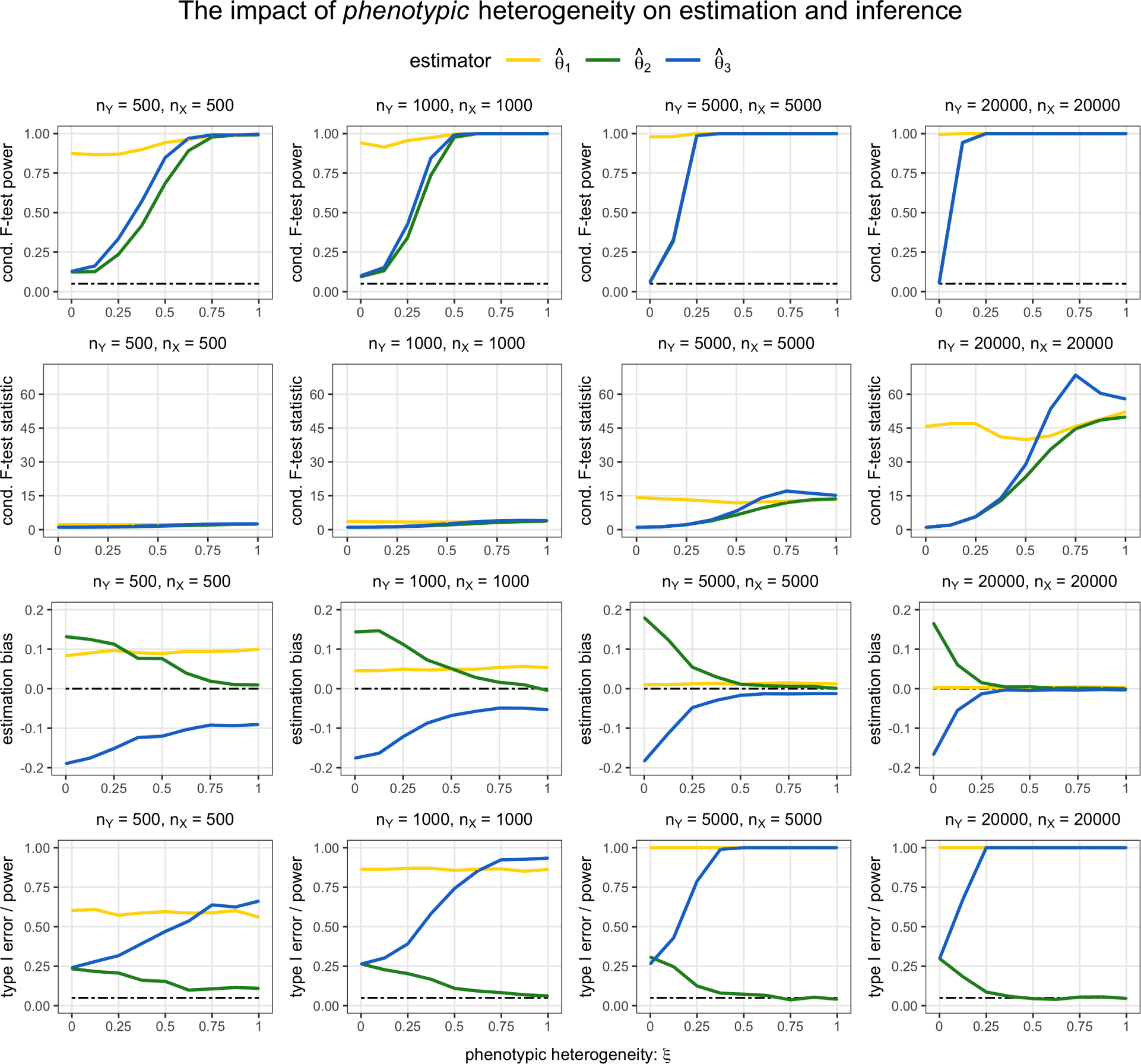
Estimation bias and inference using robust PC-GMM under varying phenotypic heterogeneity *ξ* and no overdispersion heterogeneity (*κ*^2^ = 0). The true parameter values were *θ*_1_ = *−*1*/*3, *θ*_2_ = 0, *θ*_3_ = 1/3. The nominal size of all tests was 0.05.

The results of PC-GMM estimation and inference for the case of phenotypic heterogeneity (*ξ* = 1) and varying overdispersion heterogeneity (0 *≤ κ*^2^ *≤* 1) are displayed in Figure 4. The first row displays the root-mean squared error (RMSE) performance of PC-GMM estimates, and the second row displays the type I error rates when testing the null effect *θ*_2_ = 0 as well as the power to detect the non-zero effects *θ*_1_ and *θ*_3_. When *κ*^2^ = 0, there appears to be a small loss in power going from the unrobust to robust estimates, but for larger sample sizes the robust PC-GMM method was competitive in terms of power. Importantly, however, only the robust PC-GMM method was able to control the type I error rate of *θ*_2_ near the nominal 5% level, unlike the unrobust version that had a type I error rate of nearly 40% when *κ*^2^ = 1 even in very large samples (*n_X_*= *n_Y_* = 20, 000).

**Figure 4.**
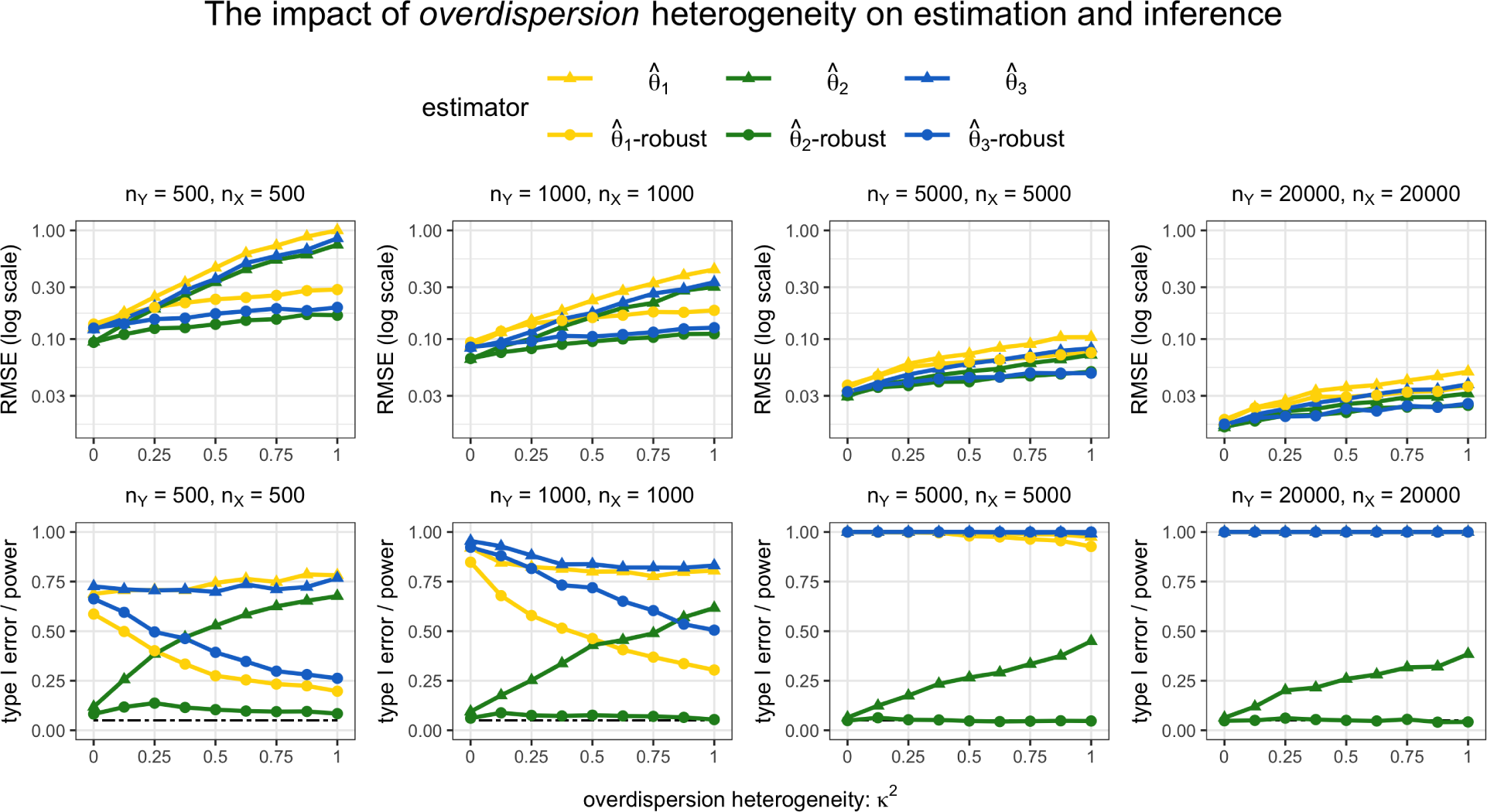
RMSE and type I error/power under phenotypic heterogeneity (*ξ* = 1) and varying overdispersion heterogeneity (0 *≤ κ*^2^ *≤* 1). The nominal size of all tests was 0.05. The true parameter values were *θ*_1_ = *−*1*/*3, *θ*_2_ = 0, *θ*_3_ = 1/3. For each risk factor *k*, 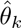-robust indicates the results using robust PC-GMM, and 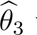 indicates the results using unrobust PC-GMM (which assumes *κ*^2^ = 0).

Strangely, in smaller samples, the methods were unable to detect the negative effect *θ*_1_ = *−*1/3 quite as well as the positive effect *θ*_3_ = 1/3. This power asymmetry has recently been discussed in the context of two-stage least squares estimates (Keane and Neal, 2023), and our findings suggest a similar phenomenon may exist here for summary data PC-GMM estimates. The robust PC-GMM method also offered improved estimation in smaller samples in terms of RMSE compared with the unrobust version when *κ*^2^ *>* 0. However, the RMSE from both methods fall with larger sample sizes. This is a direct consequence of our modelling assumptions, which imply that even the unrobust estimates should be consistent; the problem of overdispersion heterogeneity affects inference rather than estimation.

Finally, we note that the results when choosing the number of principal components to explain 99.9% of variation in the weighted genetic correlation matrix exhibited very similar patterns to those using the 99.99% threshold that are shown in Figures 3 and 4. In practice, we recommend using a fairly large number of principal components in order to reliably estimate an overdispersion parameter *κ*^2^, and hence to obtain accurate inferences. However, we should also be wary that the inclusion of principal components that are only weakly associated with the risk factors would lower the values of conditional F-statistics.

### Colocalization for BMI and T2D at the *GLP1R* gene region

Colocalization did not provide evidence supporting a shared causal variant for both BMI and T2D at the *GLP1R* locus. At the default prior values, the posterior probability of the shared causal variant hypothesis (*H*_4_) was 0.1%. The posterior probability that there is a causal variant for T2D but not for BMI (*H*_2_) was 73.3%, and the posterior probability that there are distinct causal variants for BMI and T2D (*H*_3_) was 24.4%. The posterior probability that there are distinct causal variants for BMI and T2D conditional on both variants having a causal variant (*H*_3/_(*H*_3_ + *H*_4_)) was 99.6%. This suggests the presence of phenotypic heterogeneity at this locus. A sensitivity analysis indicated little support for the shared causal variant hypothesis at any value of the *p*_12_ parameter (Supplementary Figure S2).

The conditional F-statistic relating to BMI using 40 principal components of 1088 genetic variants was *F_BMI|T_* _2_*_D_* = 2.371, while the corresponding conditional F-statistic relating to T2D risk was *F_T_* _2_*_D|BMI_* = 3.028. Conditional F-tests rejected the null hypothesis of no phenotypic heterogeneity (*P <* 0.001). In Supplementary Table 2, we also note Mendelian randomization estimates when using a fewer number of relevant principal components as instruments, which led to larger conditional F-statistics for both risk factors. For example, *F_BMI|T_* _2_*_D_* = 7.027 and *F_T_* _2_*_D|BMI_* = 18.081 for the case where only 4 relevant principal components were used.

### Mendelian randomization analyses for BMI and T2D on CAD risk

Results from the robust PC-GMM method for the analysis including BMI and T2D risk are presented in Table 1. When using 40 principal components that explained 99.9% of weighted variation in 1088 genetic variants, genetically-predicted BMI was associated with CAD risk (log odds ratio estimate per 1 standard deviation increase in BMI 1.470, 95% confidence interval [CI] 0.621, 2.319), but genetically-predicted T2D liability was not (log odds ratio estimate per 1 unit increase in the log odds of T2D risk −0.051, 95% CI −0.239, 0.136).

**Table 1.** Robust PC-GMM results: Genetically-predicted multivariable BMI and T2D effects on CAD risk using 40 principal components that explain 99.9% of weighted genetic variation in *GLP1R*.

|  | estimate | 95% CI | p-value | # PCs | # genetic variants |
| --- | --- | --- | --- | --- | --- |
| BMI | 1.470 | 0.621, 2.319 | 0.001 | 40 | 1088 |
| T2D | -0.051 | -0.239, 0.136 | 0.591 |  |  |

Estimates were similar when not accounting for overdispersed heterogeneity in the genetic associations (Supplementary Table S1), but the heterogeneity test was rejected (*P* = 0.003). The results were not very sensitive to the number of principal components included in the analysis; when using principal components explaining 95% and 99% of weighted genetic variation, estimates were similar in magnitude, but less precise (Supplementary Table S1). Finally, the results were also similar when using a fewer number of relevant principal components (*P <* 0.05 association with BMI or T2D) with larger associated conditional F-statistics (Supplementary Table S2).

### Mendelian randomization analyses for tissue-specific *GLP1R* expression on CAD risk

Results from the robust PC-GMM method for the analyses of gene expression in different tissues are presented in Figure 5 when using 21 principal components that explained 99.9% of weighted variation in 851 genetic variants. In the limited analysis comparing gene expression in pancreas, brain-caudate, and heart-atrial appendage, genetically-predicted gene expression in brain-caudate was associated with CAD risk (estimate −0.067, 95% CI −0.118, −0.015), whereas gene expression in the other tissues was not (Figure 5A). The conditional F-statistics relating to the 3 tissues were between 1.256 and 1.441. Results were similar but less precise with fewer principal components (Supplementary Table S3). Similarly, in the extended analysis, only gene expression in brain-caudate was associated with CAD risk at a conventional level of statistical significance (Figure 5B; estimate −0.108, 95% CI −0.216, −0.000). However, conditional F-statistics relating to 9 of the 10 tissues were less than 1, and significance for other tissues may have been masked by the complexity of the model.

**Figure 5.**
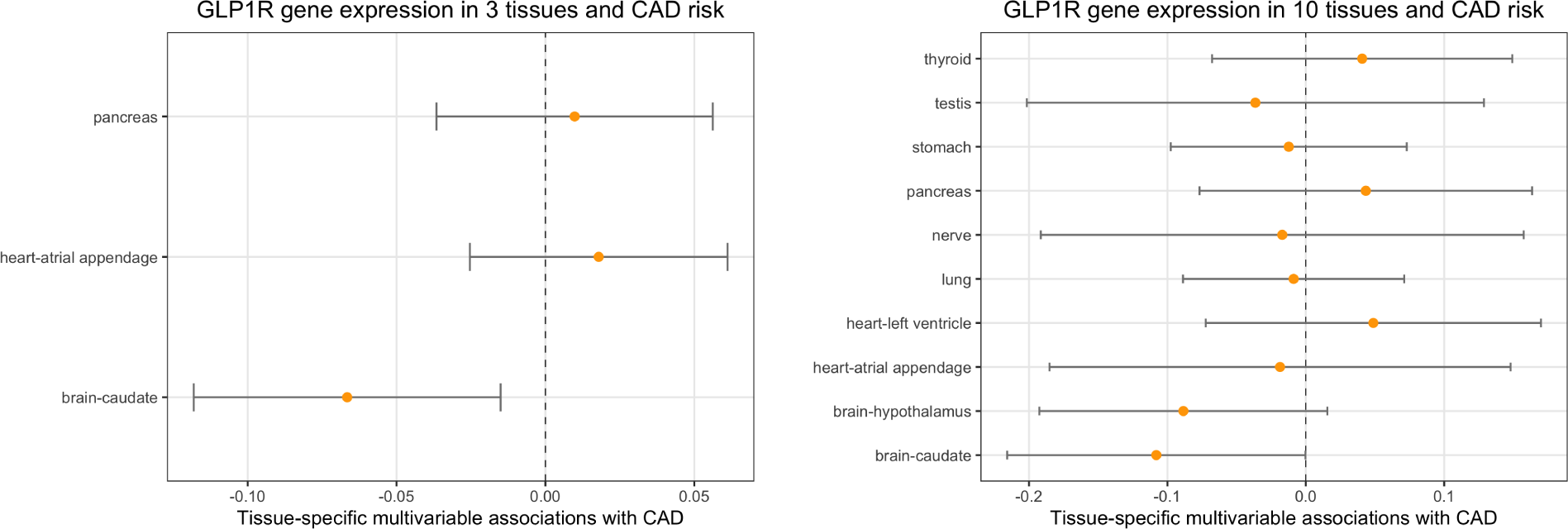
Robust PC-GMM results: Genetically-predicted multivariable effects of 3 tissues (left; Figure 5A) and 10 tissues (right; Figure 5B) on CAD risk, using principal components than explain 99.9% of weighted genetic variation in *GLP1R*.

The MR-BMA method, which compares evidence for models containing different combinations of risk factors, indicated a clear preference for the model containing brain-caudate and no other tissue (Table 2). The posterior probability for this model was 45.1%. The next highest ranking models contained testis only (posterior probability 11.0%), lung only (10.7%), nerve only (6.7%), and then stomach only (6.4%). Overall, the marginal inclusion probability for brain-caudate was 47.1%, indicating that brain-caudate was selected as a causal risk factor in models with a total posterior probability of 47.1%. The next ranking tissues by marginal inclusion probability were testis (11.8%) and lung (11.2%).

**Table 2.** MR-BMA results: Top 5 ranked models and corresponding posterior probabilities.

| model rank | tissues selected by MR-BMA | posterior probability (%) |
| --- | --- | --- |
| 1 | brain-caudate | 45.1 |
| 2 | testis | 11.0 |
| 3 | lung | 10.7 |
| 4 | nerve | 6.7 |
| 5 | stomach | 6.4 |

## Discussion

In this paper, we have developed a novel extension to multivariable *cis*-Mendelian randomization that offers accurate confidence intervals when overdispersion heterogeneity in dimension-reduced genetic associations is detected. Conventional standard errors that ignore the presence of overdispersion heterogeneity may lead to inflated type I error rates, as illustrated in our simulation study.

We demonstrated the use of this method to better understand the effect of GLP1R agonism on CAD risk. We found evidence for an association between genetically-predicted BMI and CAD risk in a multivariable model including both BMI and T2D, suggesting that the mechanistic pathway from *GLP1R* to CAD risk passes predominantly via BMI rather than T2D. We also demonstrated an association between genetically-predicted *GLP1R* gene expression in the brain and CAD risk in a multivariable model including *GLP1R* gene expression in specific areas of the brain and heart, and the pancreas. This was further validated in a multivariable model including gene expression in a wide range of tissues.

Our findings provide mechanistic insight into the effects of GLP1R agonists in preventing CAD. The results are consistent with the bodyweight lowering effects of GLP1R agonism being involved in mediating reduced CAD risk more than T2D liability reducing effects. Further, bodyweight reduction may in itself be reducing T2D liability (Gill et al., 2021). However, it is important to appreciate that reduced inflammation, blood pressure, and triglycerides are other mechanisms that are also reduced by GLP1R agonists and which may contribute to reduce CAD risk (Rakipovski et al., 2018).

These results of this study are hypothesis-generating. It should, however, be noted that these data directly relate to risk of CAD incidence, rather than progression. Thus, our results support that GLP1R agonism is likely to be efficacious through these mechanisms for CAD prevention. Further work may investigate related cardiovascular outcomes, including stroke and heart failure.

We note that our method is widely applicable for robust inference in general multivariable *cis*-Mendelian randomization investigations. For many drug targets, the mechanism of action of the drug is unclear. Multivariable MR can provide insights into plausible risk factors, which represent causal agents or surrogate markers indicating the relevant causal pathway (Schmidt et al., 2021).

In the two-sample setting considered in the simulation, there was some type I error inflation at low levels of phenotypic heterogeneity, particularly with small sample sizes and low conditional F statistics. In practice, we would advise comparing results with fewer genetic principal components (say, variants explaining 95% or 99% of variation in the weighted genetic correlation matrix), which would typically result in lower power but less bias in estimates, and with more principal components (say, variants explaining 99.9% of variation), which would typically result in greater power, but could lead to bias from weak instruments. In a one-sample setting, greater care to avoid using weak instruments would be advised. In our applied examples, results were broadly similar with different numbers of principal components.

The key strength of our method is that it offers a way to disentangle and obtain robust inferences on effects of several correlated risk factors on an outcome, thereby illuminating causal mechanisms that could potentially inform the design of therapeutic interventions. The general advantages of many summary data Mendelian randomization investigations also apply; the analyses may be less vulnerable to biases due to reverse causality, and the increasing availability of large-scale genetic association data and genotyped biobank data makes the use of such approaches for gaining insight into drug target effects both time and cost efficient.

There are, however, some limitations of our investigation. In terms of methodology, while the overdispersion correction accounts for the extra uncertainty due to random heterogeneity in genetic associations, it does not account for more idiosyncratic heterogeneity which could lead to biased estimation. In our application, we do not use cell-specific gene expression data which could strengthen causal claims and identify cell-type specific effects (Haglund et al., 2022). Moreover, the tissues we considered for our analyses were based on measured *GLP1R* expression levels, but they may not necessarily be the tissues most biologically relevant for the study of GLP1R agonism. Specifically, for *GLP1R* expression in the brain it is difficult to map these tissues onto effects, as they may be contributing to weight loss, but could also be involved in glycemia, inflammation or lipid lowering. Another limitation of our analysis is that it does not consider time varying exposure effects; previous studies have suggested that the effect of BMI on CAD risk may depend on BMI over life course (Richardson et al., 2020). It may also be useful to consider additional relevant effects of GLP1R agonism, such as reduced inflammation, blood pressure, and triglyceride levels, in order to further understand the mechanisms by which GLP1 agonists exert their effects on reducing CAD risk.

As a final semantic point, we note that the term “phenotypic heterogeneity” is used inconsistently in the literature: in some cases, it refers to distinct effects of the same genetic polymorphism (Wolf, 1997; particular for highly disruptive variants); whereas elsewhere, it refers to distinct effects of different polymorphisms in the same gene region (Nussbaum et al., 2007). We here use the latter definition, and encourage future researchers to clearly define the term in context to clarify whether heterogeneity is being explored at the variant or gene level.

In conclusion, we have presented evidence suggesting that the effect of GLP1R agonism on CAD risk operates predominantly via BMI reduction, as compared to reduced T2D liability. Separate tissue-specific analyses suggest brain mechanisms may be relevant for CAD risk compared with other considered tissues. We hope our investigation serves as both a substantive analysis and a didactic example of how phenotypic heterogeneity at a gene corresponding to a druggable target can be used to investigate the mechanism and site-of-action of the causal effect of pharmacological intervention of that target on a disease outcome.

## Data Availability

The data that support the findings of this study are openly available from UK Biobank at http://www.nealelab.is/uk-biobank; the DIAMANTE consortium at https://kp4cd.org/node/169; the GIANT consortium at https://portals.broadinstitute.org/collaboration/giant/index.php/GIANT_consortium; the Genotype-Tissue Expression (GTEx) project version 8 at https://gtexportal.org/home/; and the CARDIoGRAMplusC4D consortium at http://www.cardiogramplusc4d.org/.

https://kp4cd.org/node/169

https://portals.broadinstitute.org/collaboration/giant/index.php/GIANT_consortium

https://gtexportal.org/home/

http://www.cardiogramplusc4d.org/

