## Supplementary Material for "Robust use of phenotypic heterogeneity at drug target genes for mechanistic insights: application of *cis*-multivariable Mendelian randomization to *GLP1R* gene region"

### The robust PC-GMM method

#### Linear model with dimension-reduced instruments

Let $X = (X_1, \ldots, X_K)'$ denote a $K$-vector of risk factors, $Y$ an outcome, and $Z = (Z_1, \ldots, Z_M)'$ an $M$-vector of genetic variants. Our focus is on a *cis*-gene analysis where genetic variants from a single region are in highly structured correlation. Let $\Lambda$ denote the $M \times L$ matrix where its columns are the first $L$ principal components ($K \leq L < M$) of a weighted sample correlation matrix of genetic variants.[1] We consider the following linear IV model with homoscedastic errors

$$Y = \omega + \theta_0' X + \alpha'(\Lambda' Z) + U \quad (1)$$

$$X = \psi + \gamma'(\Lambda' Z) + V \quad (2)$$

where $E[U|\Lambda' Z] = 0$, $E[V|\Lambda' Z] = 0$, $E[U^2|\Lambda' Z] = \sigma_U^2$, $E[VV'|\Lambda' Z] = \Sigma_V$, and $(\omega, \psi, \gamma, \theta_0)$ are unknown parameters. We assume $\alpha$ is a mean-zero random effect which is uncorrelated with all other variables. We are interested in estimation and inference on the $K$-vector of risk factor effects on the outcome $\theta_0$, using only two-sample summary data that is often made publicly available.

For each variant $m$ and risk factor $k$, we have access to estimates $\widehat{\beta}_{X_{km}}$ and standard errors $\sigma_{X_{km}}$ from univariable $Z_m$ on $X_k$ linear regressions from an $n_X$-sized sample, and from a non-overlapping $n_Y$-sized sample, we observe measured associations $(\widehat{\beta}_{Y_m}, \sigma_{Y_m})$ from univariable $Z_m$ on $Y$ linear regressions. Both random samples are drawn from the joint distribution of $(Y, X, Z)$. We assume knowledge of an $M \times M$ genetic correlation (or linkage disequilibrium) matrix $\rho$, where its $(m_1, m_2)$-th element $\rho_{m_1 m_2}$ denotes the correlation between the $m_1$-th and $m_2$-th genetic variant. Finally, we also assume knowledge of a $K \times K$ risk factor correlation matrix $\tau$, where its $(k_1, k_2)$-th element $\tau_{k_1 k_2}$ denotes the correlation between $X_{k_1}$ and $X_{k_2}$.

Substituting (2) into (1), we have $Y = (\omega + \theta_0'\psi) + (\gamma\theta_0 + \alpha)'(\Lambda' Z) + (U + \theta_0' V)$. Thus, $Cov(\Lambda' Z, Y) = Var(\Lambda' Z)(\gamma\theta_0 + \alpha)$, which leads to a model

$$\Gamma = \gamma\theta_0 + \alpha, \text{ where } \alpha \sim N(0_{L\times 1}, I_L \kappa^2 n_Y^{-1}) \quad (3)$$

where $\Gamma$ is the $L$-vector of coefficients from a population multivariable regression of $\Lambda'Z$ on $Y$, and $\gamma$ is the $L \times K$ matrix such that its $k$-th column $\gamma_k$ is the $L$-vector of coefficients from a population multivariable regression of $\Lambda'Z$ on $X_k$. The random effects $\alpha$ are mutually uncorrelated, and assumed to be normally distributed scaled up to an unknown overdispersion variance parameter $\kappa^2$.

**Proposition 1** (Two-sample summary data associations). *Using the two-sample univariable summary data described above, we can construct multivariable estimates* $(\widehat{\Gamma}, \widehat{\Sigma}_\Gamma)$ *and* $(vec(\widehat{\gamma}), \widehat{\Sigma}_\gamma)$, *such that*

$$\begin{bmatrix} \sqrt{n_Y}(\widehat{\Gamma} - \gamma\theta_0) \\ \sqrt{n_X}(vec(\widehat{\gamma}) - vec(\gamma)) \end{bmatrix} \xrightarrow{D} N\left( \begin{bmatrix} 0_{L\times 1} \\ 0_{LK\times 1} \end{bmatrix}, \begin{bmatrix} \Sigma_\Gamma + I_L\kappa^2 & 0_{L\times LK} \\ 0_{LK\times L} & \Sigma_\gamma \end{bmatrix} \right),$$

$\widehat{\Sigma}_\Gamma - \Sigma_\Gamma \xrightarrow{P} 0$, *and* $\widehat{\Sigma}_\gamma - \Sigma_\gamma \xrightarrow{P} 0$, *as* $(n_X, n_Y) \to \infty$.

#### Conditional F-statistics

We require some additional notation to construct conditional F-statistics in our setting of dimension-reduced genetic associations. Let $\widehat{\Sigma}_{\gamma,k}$ denote the matrix $\widehat{\Sigma}_\gamma$ that is rearranged such that the $k$-th column is moved left to be the first column, and the $k$-th row is moved up to be the first row. Let $\widehat{\gamma}_{-k}$ denote the $L \times (K-1)$ matrix equal to $\widehat{\gamma}$ without the $k$-th column, let $\gamma_{-k}$ denote the $L \times (K-1)$ matrix equal to $\gamma$ without the $k$-th column, and let $h(\delta) = [I_L \quad -I_L \otimes \delta']$ denote the $L \times LK$ matrix where $\delta$ is a $(K-1)$-vector of unknown parameters, and where $\otimes$ denotes the Kronecker product.

Then, for any risk factor $k$, we consider conditional F-statistics[2] given by

$$F_{k|-k} = \frac{n_X}{L-K+1} \min_\delta \{(\widehat{\gamma}_k - \widehat{\gamma}_{-k}\delta)'[h(\delta)\widehat{\Sigma}_{\gamma,k}h(\delta)']^{-1}(\widehat{\gamma}_k - \widehat{\gamma}_{-k}\delta)\}.$$

**Proposition 2** (Conditional F-statistics). *For any risk factor $k$, under the null hypothesis $H_{0k}$ : $\gamma_k - \gamma_{-k}\delta = 0$ uniquely at some $\delta = \delta_0$, $F_{k|-k}(L-K+1) \xrightarrow{D} \chi^2_{L-K+1}$ as $n_X \to \infty$.*

Under the null hypothesis $H_{0k} : \gamma_k - \gamma_{-k}\delta = 0$ uniquely at some $\delta = \delta_0$, Proposition 2 shows that the statistic $F_{k|-k}(L-K+1)$ should behave like a $\chi^2_{L-K+1}$ random variable in large samples. This allows us to perform a test of no phenotypic heterogeneity for any risk factor $k$, with a rejection of the null hypothesis $H_{0k}$ suggesting evidence for phenotypic heterogeneity.

#### Robust PC-GMM: 2-step estimation of $\theta_0$ and $\kappa^2$

By Proposition 1, we have $E[\widehat{\Gamma} - \widehat{\gamma}\theta_0] = 0_{L\times 1}$, so that there are $L$ moment equations which describe $\theta_0$. To identify $\theta_0$, we require the usual rank condition that the column rank of $\gamma$ is at least $K$.

Let $\widehat{g}(\theta) = \widehat{\Gamma} - \widehat{\gamma}\theta$ denote an $L$-vector of estimating equations. Then, using Proposition 1, an estimator of the variance of $\widehat{g}(\theta_0)$ is given by $\widehat{\Omega}(\theta_0, \kappa^2) = n_Y^{-1}(\widehat{\Sigma}_\Gamma + I_L\kappa^2) + n_X^{-1}\varphi(\theta_0)\widehat{\Sigma}_\gamma\varphi(\theta_0)'$, where $\varphi(\theta) = \theta' \otimes I_L$. Thus, under knowledge of $\kappa^2$, we consider $\widehat{Q}(\theta, \kappa) = \widehat{g}(\theta)'\widehat{\Omega}(\theta, \kappa^2)^{-1}\widehat{g}(\theta)$ as the oracle continuously-updating GMM[3,4] criterion function based on the optimal weighting matrix.

**Proposition 3** (Oracle GMM estimation)**.** *Under regularity conditions, the oracle GMM estimator* $\bar{\theta} = \arg\min_\theta \widehat{Q}(\theta, \kappa^2)$ *is consistent for* $\theta_0$*, and is asymptotically distributed* $\sqrt{n_Y}(\bar{\theta} - \theta_0) \overset{D}{\to} N(0_{K\times 1}, \Sigma_\theta)$ *where* $\Sigma_\theta = (\gamma'\Omega^{-1}\gamma)^{-1}$ *where* $\Omega = \Sigma_\Gamma + I_L\kappa^2 + c\varphi(\theta_0)\Sigma_\gamma\varphi(\theta_0)'$ *and* $n_X^{-1}n_Y \to c$*, as* $(n_X, n_Y) \to \infty$*.*

Proposition 3 implies that the standard errors of $\bar{\theta}$ need to account for the extra uncertainty due to the random direct effects, which is represented by the overdispersion variance parameter $\kappa^2$. However, consistent estimation of $\theta_0$ is still possible by ignoring the extra uncertainty. In particular, $\widehat{\theta}_1 = \arg\min_\theta \widehat{Q}(\theta, 0)$ is a consistent GMM estimator of $\theta$. We can then use this preliminary estimator $\widehat{\theta}_1$ to pin down $\kappa^2$.

Noting that $\widehat{Q}(\widehat{\theta}_1, \kappa^2) \overset{D}{\to} \chi^2_{L-K}$, an estimator $\widehat{\kappa}^2$ of $\kappa^2$ solves the estimating equation $\widehat{Q}(\widehat{\theta}_1, \widehat{\kappa}^2) - (L - K) = 0$. Then, the 2-step estimator of $\theta_0$ is given by $\widehat{\theta} = \arg\min_\theta \widehat{Q}(\theta, \widehat{\kappa}^2)$. Finally, it is straightforward to estimate the standard errors of $\hat{\theta}$ based on Proposition 3. For the $k$-th risk factor, the estimated standard errors are given by the square root of the $(k, k)$-th element of the matrix $(\widehat{\gamma}'\widehat{\Omega}(\widehat{\theta}, \widehat{\kappa}^2)^{-1}\widehat{\gamma})^{-1}$.

Finally, for the unrobust version of the PC-GMM method, a test of overidentifying restrictions[5] (henceforth, heterogeneity test) is a useful way to assess the coherency of evidence over all instruments. Assuming that $\theta_0$ is identified (so that there are at least $K$ distinct valid instruments in the vector $\Lambda'Z$), if one of the instruments is invalid, then $\widehat{g}(\theta_0)$ no longer has mean zero, and therefore we would expect the GMM criterion $\widehat{Q}(\theta_0, 0)$ to deviate further away from 0. This is the intuition behind the heterogeneity test.

More formally, by very similar arguments used in Proof of Proposition 2, it can be shown under no overdispersion heterogeneity ($\kappa^2 = 0$) that $\min_\theta \widehat{Q}(\theta, 0) \overset{D}{\to} \chi^2_{L-K}$. Therefore, we can compute a heterogeneity test by comparing the statistic $\widehat{Q}(\widehat{\theta}, 0)$ against a relevant critical value from the $\chi^2_{L-K}$ distribution. Let $c_\alpha$ denote the $(1-\alpha)$-th quantile of the $\chi^2_{L-K}$ distribution. If $\widehat{Q}(\widehat{\theta}, 0) > c_\alpha$, then we reject the null hypothesis of instrument coherency for an $\alpha$-level test.

### Further empirical results

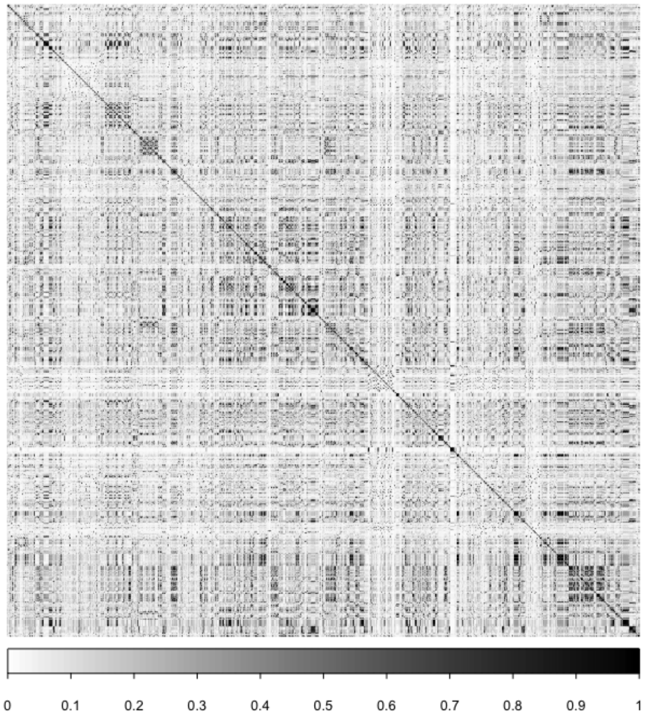


Supplementary Figure S1. The absolute values of the correlation matrix of 851 genetic variants in *GLP1R*.

| PC-GMM method | risk factor | estimate | 95% CI | p-value | # PCs | het. test | cond. F-stat. |
|---|---|---|---|---|---|---|---|
| robust (99.9%) | BMI | 1.470 | 0.621, 2.319 | 0.001 | 40 | - | 2.371 |
| | T2D | -0.051 | -0.239, 0.136 | 0.591 | | | 3.028 |
| unrobust (99.9%) | BMI | 1.484 | 0.828, 2.140 | <0.001 | | 0.003 | - |
| | T2D | -0.139 | -0.288, 0.010 | 0.068 | | | - |
| robust (99%) | BMI | 2.876 | 0.618, 5.135 | 0.013 | 17 | - | 3.271 |
| | T2D | -0.094 | -0.417, 0.229 | 0.569 | | | 4.476 |
| unrobust (99%) | BMI | 3.185 | 2.023, 4.348 | <0.001 | | 0.001 | - |
| | T2D | -0.293 | -0.527, -0.060 | 0.014 | | | - |
| robust (95%) | BMI | 0.213 | -3.538, 3.964 | 0.911 | 8 | - | 4.100 |
| | T2D | 0.286 | -0.730, 1.301 | 0.582 | | | 5.182 |
| unrobust (95%) | BMI | 4.260 | 2.499, 6.020 | <0.001 | | <0.001 | - |
| | T2D | -0.522 | -0.952 -0.093 | 0.017 | | | - |

Supplementary Table S1. PC-GMM results: Genetically-predicted multivariable BMI and T2D effects on CAD risk. The percentages next to the PC-GMM method indicate the percentage of weighted genetic variation explained in *GLP1R* by the number of principal components used.

| # PCs | risk factor | estimate | 95% CI | p-value | cond. F-stat. |
|---|---|---|---|---|---|
| 4 | BMI | 2.966 | 0.903, 5.030 | 0.005 | 7.027 |
| | T2D | -0.092 | -0.410, 0.226 | 0.570 | 18.081 |
| 6 | BMI | 2.421 | 0.878, 3.964 | 0.002 | 6.086 |
| | T2D | -0.045 | -0.305, 0.214 | 0.731 | 10.847 |
| 8 | BMI | 1.978 | 0.682, 3.273 | 0.003 | 5.285 |
| | T2D | -0.063 | -0.307, 0.180 | 0.610 | 7.464 |
| 10 | BMI | 0.873 | -0.391, 2.136 | 0.176 | 4.806 |
| | T2D | -0.012 | -0.265, 0.242 | 0.929 | 4.692 |
| 12 | BMI | 1.290 | 0.112, 2.468 | 0.032 | 3.961 |
| | T2D | -0.037 | -0.251, 0.178 | 0.739 | 5.393 |
| 14 | BMI | 1.427 | 0.281, 2.572 | 0.015 | 3.404 |
| | T2D | -0.007 | -0.208, 0.193 | 0.944 | 5.498 |

Supplementary Table S2. Robust PC-GMM results: Genetically-predicted tissue-specific multivariable effects on CAD risk using a given number of relevant principal components ($P<0.05$ association with BMI and T2D) that maximise the minimum conditional F-statistic over both traits.

| var. expl. | tissue | estimate | 95% CI | p-value | # PCs |
|---|---|---|---|---|---|
| 99.9% | brain-caudate | -0.067 | -0.118, -0.015 | 0.011 | 21 |
| | heart-atrial appendage | 0.018 | -0.025, 0.061 | 0.418 | |
| | pancreas | 0.010 | -0.037, 0.056 | 0.678 | |
| 99% | brain-caudate | -0.097 | -0.163, -0.031 | 0.004 | 10 |
| | heart-atrial appendage | 0.029 | -0.014, 0.072 | 0.191 | |
| | pancreas | 0.005 | -0.029, 0.039 | 0.790 | |
| 95% | brain-caudate | -0.130 | -0.271, 0.012 | 0.072 | 5 |
| | heart-atrial appendage | 0.040 | -0.032, 0.112 | 0.277 | |
| | pancreas | 0.004 | -0.041, 0.049 | 0.859 | |

Supplementary Table S3. Robust PC-GMM results: Genetically-predicted tissue-specific multivariable effects on CAD risk using *GLP1R* variants.

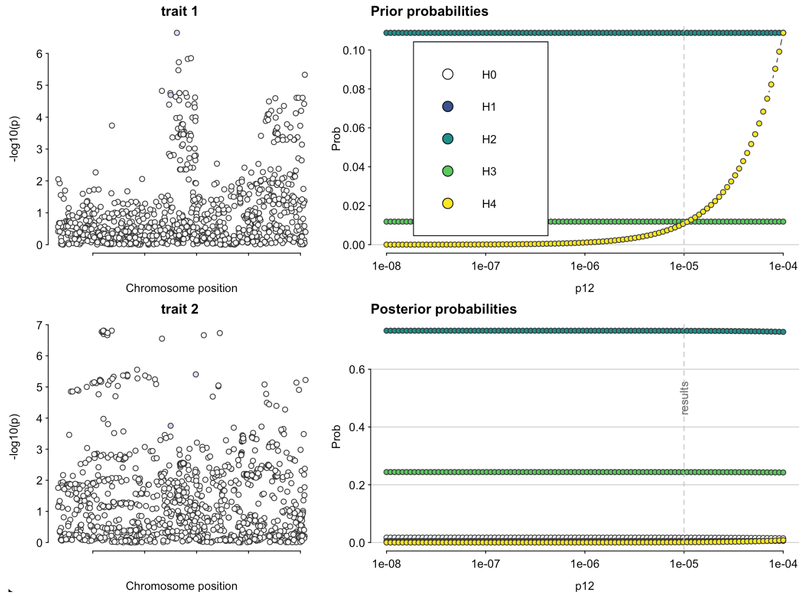


Supplementary Figure S2. Sensitivity of coloc results to the choice of prior $p_{12}$ on the shared causal variant hypothesis.

### Further simulation results

We consider the impact of mis-specifying the true trait correlations $\rho$ in the same simulation design discussed in the main text, with $\xi = 1$ (phenotypic heterogeneity; all 3 risk factors have 5 distinct causal variants) and $\kappa^2 = 0.5$ (moderate overdispersion heterogeneity). Supplementary Figure S3 shows the impact on type I error (relating to $\theta_2 = 0$) and power (relating to $\theta_1 = -1/3$ and $\theta_3 = 1/3$) of specifying trait correlations $\widehat{\rho}$ instead of $\rho$.

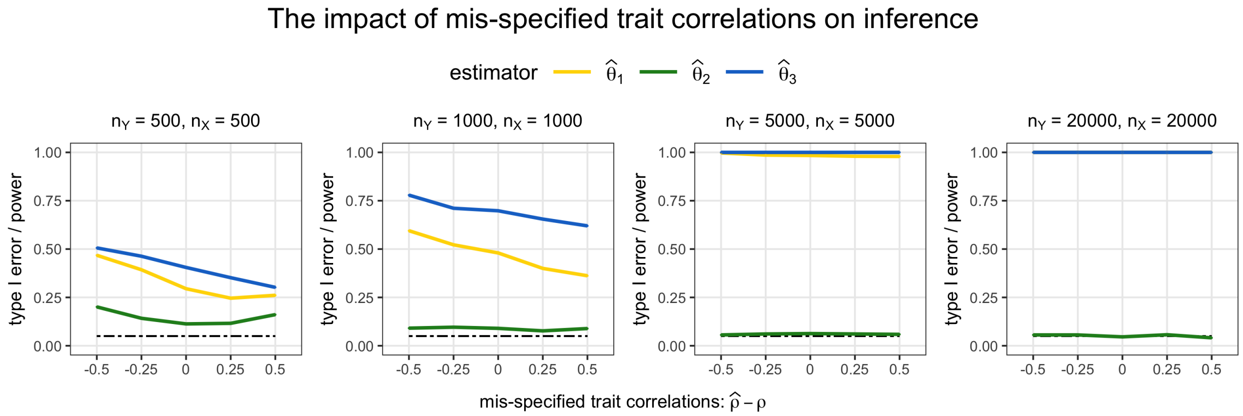


Supplementary Figure S3. Type I error/power varying with mis-specification of trait correlations. The true causal effect is $(\theta_1, \theta_2, \theta_3)' = (-1/3, 0, 1/3)'$.

From Supplementary Figure S3, we find that the results were not sensitive to mis-specification of trait correlations for large enough sample sizes. For small samples ($n_X = n_Y = 500$), mis-specification of trait correlations does appear to harm an already inflated type I error rate. Interestingly, the specified trait correlations also appear to impact the power properties: under-estimating trait correlations ($\widehat{\rho} < \rho$) appeared to be less harmful in terms of power compared with over-estimating the correlations. Although, more generally, the impact on power may depend on several other model parameters, such as the direction of the true trait correlations $\rho$ and the direction of the causal effect $\theta = (\theta_1, \theta_2, \theta_3)'$.

### Appendix

#### Proof of Proposition 1

##### Asymptotic distributions of $\widehat{\Gamma}$ and $\widehat{\gamma}$

From Equations (1)–(3), we have the reduced form outcome model $Y = \eta + (\gamma\theta_0 + \alpha)'(\Lambda' Z) + \varepsilon$, where $\eta = \omega + \theta_0'\psi$, and $\varepsilon = U + \theta_0' V$, so that $E[\varepsilon|\Lambda' Z] = 0$ and $var(\varepsilon|\Lambda' Z) = \sigma_\varepsilon^2$.

Let $\widehat{\Gamma} = \widehat{var}(\Lambda' Z)^{-1}\widehat{cov}(\Lambda' Z, Y)$ be the estimated coefficient of a multivariable linear regression of $Y$ on $\Lambda' Z$ with a constant term included. Note that $E[\widehat{\Gamma}] = \gamma\theta_0$, $\sqrt{n_Y}\big(\widehat{cov}(\Lambda' Z, Y) - cov(\Lambda' Z, Y)\big) \overset{D}{\to} N(0_{L\times 1}, var(\Lambda' Z)\sigma_\varepsilon^2 + var(\Lambda' Z) I_L \kappa^2 var(\Lambda' Z)')$ by a central limit theorem (CLT), and $\widehat{var}(\Lambda' Z) \overset{P}{\to} var(\Lambda' Z)$ by the weak law of large numbers (WLLN).

Hence, by Cramer's theorem, $\sqrt{n_Y}(\widehat{\Gamma} - \gamma\theta_0) \overset{D}{\to} N(0_{L\times 1}, I_L\kappa^2 + \Sigma_\Gamma)$ where $\Sigma_\Gamma = var(\Lambda' Z)^{-1}\sigma_\varepsilon^2$, and $\sigma_\varepsilon^2 = var(Y) - cov(\Lambda' Z, Y)' var(\Lambda' Z)^{-1} cov(\Lambda' Z, Y)$.

For each risk factor $k = 1, \ldots, K$, let $\widehat{\gamma}_k = \widehat{var}(\Lambda' Z)^{-1}\widehat{cov}(\Lambda' Z, X_k)$ be the estimated coefficients of a multivariable regression of $X_k$ on $\Lambda' Z$ with a constant term included. By similar arguments to the above, we have $\sqrt{n_X}(\widehat{\gamma}_k - \gamma_k) = var(\Lambda' Z)^{-1}\sqrt{n_X}\widehat{cov}(\Lambda' Z, V_k) + o_P(1)$. Therefore, by a CLT and the Cramer-Wold device, $\sqrt{n_X}(vec(\widehat{\gamma}) - vec(\gamma)) \overset{D}{\to} N\big(0_{LK\times 1}, \Sigma_\gamma\big)$, where $\Sigma_\gamma$ is a $LK \times LK$ variance–covariance matrix.

Under homoscedastic errors, $\Sigma_\gamma$ has the following block structure

$$\Sigma_\gamma = \begin{bmatrix} \underset{(L\times L)}{\Sigma_{\gamma,11}} & \underset{(L\times L)}{\Sigma_{\gamma,12}} & . & . & \underset{(L\times L)}{\Sigma_{\gamma,1K}} \\ \underset{(L\times L)}{\Sigma_{\gamma,21}} & \underset{(L\times L)}{\Sigma_{\gamma,22}} & & & . \\ . & & . & & . \\ . & & & . & . \\ \underset{(L\times L)}{\Sigma_{\gamma,K1}} & . & . & . & \underset{(L\times L)}{\Sigma_{\gamma,KK}} \end{bmatrix},$$

where $\Sigma_{\gamma,k_1k_2} = \big(var(\Lambda' Z)^{-1}\big(cov(X_k, X_m) - cov(\Lambda' Z, X_k)' var(\Lambda' Z)^{-1} cov(\Lambda' Z, X_m)\big)\big)$ for any risk factors $k_1$ and $k_2$.

**Constructing $(\widehat{\Gamma}, \widehat{\Sigma}_\Gamma)$ and $(\widehat{\gamma}, \widehat{\Sigma}_\gamma)$ from two-sample summary data**

We follow the strategy of Wang and Kang (2022).[6] First, we need to calculate principal components of $\widehat{cov}(Z)$. For each variant $m$, we have $(n_Y \sigma^2_{Y_m} + \widehat{\beta}^2_{Y_m})^{-1} = \widehat{var}(Y)^{-1}\widehat{var}(Z_m)$. Let $A_Y$ be the $M \times M$ matrix with its $(m_1, m_2)$-th element given by $\rho_{m_1 m_2}(n_Y \sigma^2_{Y_{m_1}} + \widehat{\beta}^2_{Y_{m_1}})^{-\frac{1}{2}}(n_Y \sigma^2_{Y_{m_2}} + \widehat{\beta}^2_{Y_{m_2}})^{-\frac{1}{2}}$, so that $A_Y = \widehat{var}(Y)^{-1}\widehat{var}(Z)$. Let $\Lambda$ be the $M \times L$ matrix with its columns given by first $L$ principal components of $A_Y$.

Let $b_{Y_m} = (n_Y \sigma^2_{Y_m} + \widehat{\beta}^2_{Y_m})^{-1}\widehat{\beta}_{Y_m} = \widehat{var}(Y)^{-1}\widehat{cov}(Z_m, Y)$, so that $b_Y = (b_{Y_1}, \ldots, b_{Y_p})' = \widehat{cov}(Z, Y)\widehat{var}(Y)^{-1}$. Then, $\widehat{\Sigma}_\Gamma = \big(\Lambda' A_Y \Lambda\big)^{-1}\big[1 - \big(\Lambda' b_Y\big)'\big(\Lambda' A_Y \Lambda\big)^{-1}\big(\Lambda' b_Y\big)\big] \xrightarrow{P} \Sigma_\Gamma$ by WLLN. Also, note that $\widehat{\Gamma} = \big(\widehat{\Lambda}' A_Y \widehat{\Lambda}\big)^{-1}\widehat{\Lambda}' b_Y$.

We can use a similar strategy for the risk factor model. For each variant $m$ and risk factor $k$, note that $(n_X \sigma^2_{X_{km}} + \widehat{\beta}^2_{X_{km}})^{-1} = \widehat{var}(X_k)^{-1}\widehat{var}(Z_m)$. For each risk factor $k$, let $A_{X_k}$ be the $M \times M$ matrix with its $(m_1, m_2)$-th element given by $\rho_{m_1 m_2}(n_X \sigma^2_{X_{km_1}} + \widehat{\beta}^2_{X_{km_1}})^{-\frac{1}{2}}(n_X \sigma^2_{X_{km_2}} + \widehat{\beta}^2_{X_{km_2}})^{-\frac{1}{2}}$, so that $A_{X_k} = \widehat{var}(X_k)^{-1}\widehat{var}(Z)$.

Let $b_{X_{km}} = (n_X \sigma^2_{X_{km}} + \widehat{\beta}^2_{X_{km}})^{-1}\widehat{\beta}_{X_{km}} = \widehat{var}(X_k)^{-1}\widehat{cov}(Z_m, X_k)$, so that $b_{X_k} = (b_{X_{k1}}, \ldots, b_{X_{kM}})' = \widehat{cov}(Z, X_m)\widehat{var}(X_m)^{-1}$. Then,

$$
\begin{aligned}
\widehat{\Sigma}_{\gamma,k_1k_2} &= \Big(\big(\Lambda A_{X_{k_1}}\Lambda\big)^{\frac{1}{2}}\big(\Lambda A_{X_{k_2}}\Lambda\big)^{\frac{1}{2}}\Big)^{-1}\Big[\tau_{k_1k_2} - \big(\Lambda' b_{X_{k_1}}\big)\Big(\big(\Lambda A_{X_{k_1}}\Lambda\big)^{\frac{1}{2}}\big(\Lambda A_{X_{k_2}}\Lambda\big)^{\frac{1}{2}}\Big)^{-1}\big(\Lambda' b_{X_{k_2}}\big)\Big] \\
&\xrightarrow{P} \Sigma_{\gamma,k_1k_2},
\end{aligned}
$$

where the second line follows by WLLN. Finally, $\widehat{\gamma}_k = \big(\Lambda A_{X_k}\Lambda\big)^{-1}\Lambda' b_{X_k}$, and $\widehat{\gamma} = (\widehat{\gamma}_1, \ldots, \widehat{\gamma}_K)$.

#### Proof of Proposition 2

We derive the asymptotic distribution of the statistic $\widehat{T} = F_{k|-k}(L - K + 1)$ under the null hypothesis $H_{0k} : \gamma_k - \gamma_{-k}\delta_0 = 0$ for some unique $\delta_0 \in \mathbb{R}^{K-1}$. Let $\widehat{m}_k(\delta) = \widehat{\gamma}_k - \widehat{\gamma}_{-k}\delta$, $\widehat{\Omega}_k(\delta) = h(\delta)\widehat{\Sigma}_{\gamma,k}h(\delta)'$, and $\Omega_k = h(\delta_0)\Sigma_{\gamma,k}h(\delta_0)'$. Then, under standard GMM arguments, the estimate $\widehat{\delta} = \arg\min \widehat{m}_k(\delta)'\widehat{\Omega}_k(\delta)^{-1}\widehat{m}_k(\delta)$ satisfies the first order expansion

$$\sqrt{n_X}(\widehat{\delta} - \delta_0) = -(M'_{-k}\Omega_k^{-1}M_{-k})^{-1}M'_{-k}\Omega_k^{-1}\sqrt{n_X}\widehat{m}_k(\delta_0) + o_P(1),$$

where $M_{-k} = -\gamma_{-k}$. Then, note that $\sqrt{n_X}(\widehat{m}_k(\widehat{\delta}) - \widehat{m}_k(\delta_0)) = M_{-k}\sqrt{n_X}(\widehat{\delta} - \delta_0) + o_P(1)$. Hence, for $R_k = I_L - \Omega_k^{-\frac{1}{2}}M_{-k}\big(M'_{-k}\Omega_k^{-1}M_{-k})^{-1}M'_{-k}\Omega_k^{-\frac{1}{2}}$,

$$\Omega_k^{-\frac{1}{2}}\sqrt{n_X}\widehat{m}_k(\widehat{\delta}) = R_k\Omega_k^{-\frac{1}{2}}\sqrt{n_X}\widehat{m}_k(\delta_0) + o_P(1).$$

Now, since $\widehat{T} = n_X\widehat{m}_k(\widehat{\delta})'\widehat{\Omega}_k(\delta)^{-1}\widehat{m}_k(\widehat{\delta})$, and $\widehat{T} - n_X\widehat{m}_k(\widehat{\delta})'\Omega_k^{-1}\widehat{m}_k(\widehat{\delta}) = o_P(1)$ by Proposition 1, we have $\widehat{T} = \mathcal{U}_k' R_k \mathcal{U}_k + o_P(1)$ where $\mathcal{U}_k = \Omega_k^{-\frac{1}{2}}\sqrt{n_X}\widehat{m}_k(\delta_0)$. Thus, $\widehat{T} \xrightarrow{D} \chi^2_{L-K+1}$ since $R_k$ is idempotent of rank $L-(K-1)$, and from Proposition 1, $\mathcal{U}_k \sim N(0, I_L)$ as $n_X \to \infty$.

#### Proof of Proposition 3

The oracle GMM estimator which assumes knowledge of $\kappa^2$ is given by $\bar{\theta} = \arg\min_\theta \widehat{Q}(\theta, \kappa^2)$, where $\widehat{Q}(\theta, \kappa^2) = \widehat{g}(\theta)'\widehat{\Omega}(\theta, \kappa^2)^{-1}\widehat{g}(\theta)$. Consistency of $\bar{\theta}$ for $\theta_0$ is given by standard GMM arguments; see, for example, Theorem 3.1 of Newey and McFadden (1994).[7] Moreover, by the first order condition, we have $\nabla_\theta \widehat{Q}(\bar{\theta}, \kappa^2) = 0$.

By the mean value theorem, there exists $\dot{\theta} \in \mathbb{R}^K$ on the line segment joining $\bar{\theta}$ and $\theta_0$ such that $\nabla_\theta \widehat{Q}(\theta_0, \kappa^2) + \nabla_{\theta\theta'} \widehat{Q}(\dot{\theta}, \kappa^2)(\bar{\theta} - \theta_0) = 0_{K\times 1}$. By standard GMM arguments, it can be shown that $n_Y \nabla_{\theta\theta'} \widehat{Q}(\dot{\theta}, \kappa^2) \overset{P}{\to} -(\gamma'\Omega^{-1}\gamma)^{-1}$ , where $\Omega = \Sigma_\Gamma + I_L\kappa^2 + c\varphi(\theta_0)\Sigma_\gamma\varphi(\theta_0)'$, $\varphi(\theta_0) = \theta_0' \otimes I_L$, and $\sqrt{n_Y}\nabla_\theta \widehat{Q}(\theta_0, \kappa^2) = -\gamma'\Omega^{-1}\sqrt{n_Y}\hat{g}(\theta_0) + o_P(1) \overset{D}{\to} N(0_{K\times 1}, \gamma'\Omega^{-1}\gamma)$. Therefore, by Cramer's theorem, $\sqrt{n_Y}(\bar{\theta} - \theta_0) \overset{D}{\to} N(0_{K\times 1}, (\gamma'\Omega^{-1}\gamma)^{-1})$.
